# Early time-restricted eating improves an actigraphy-estimated sleep quality in women with overweight or obesity

**DOI:** 10.1101/2025.07.15.25331590

**Authors:** Beeke Peters, Jennifer Jokisch, Julia Schwarz, Bettina Schuppelius, Andreas F. H. Pfeiffer, Andreas Michalsen, Achim Kramer, Olga Pivovarova-Ramich

## Abstract

**Background & Aims:** Metabolic disorders are tightly linked to sleep disturbances. First evidence showed that time-restricted eating (TRE), a growingly popular approach to combat cardiometabolic diseases, can also affect sleep quality. Recommendations for a preferable eating time window are pending. Our aim was to investigate the effects von early and late TRE on sleep quality in women with overweight and obesity.

**Methods:** A total of 31 women with overweight and obesity were included in the controlled randomized crossover study ChronoFast. After a 2-4-week baseline period, participants were randomly allocated to two-weeks early time-restricted eating (eTRE) or late time-restricted eating (lTRE). The phases were switched after a washout phase. During dietary interventions participants were only allowed to consume caloric foods and drinks in the restricted eating window (eTRE: 8 am toTRE 4 pm; lTRE: 1 pm to 9 pm). Assessment of sleep metrics was performed subjectively, using Pittsburgh Sleep Quality Index (PSQI) and self-report of sleep quality, and objectively, by blinded actigraphy. Hunger and satiety scores were examined using a visual analogue scale (VAS).

**Results:** While subjective sleep quality (PSQI and self-reported sleep quality) remained unchanged, an improvement of sleep efficiency (p = 0.047) and sleep fragmentation index (SFI) (p = 0.029) was observed in eTRE intervention compared to baseline as estimated by actigraphy. There were no correlations between hunger and satiety and sleep quality, with no differences of hunger and satiety scores between eTRE and lTRE assessed in the evening on the last day of each intervention.

**Conclusions:** eTRE but not lTRE improved objective sleep-quality, which was not related to the feeling of hunger and satiety. eTRE might be more effective strategy for well-being and sleep-related metabolic health outcomes.

ClincialTrials.gov number, NCT04351672 (registered on April 17, 2020).

## 1. Introduction

Sleep is important for physical and mental health as well as for quality of life and well-being [1]. However, sleep disorders are among the most common health complaints in the population. Insomnia or (respectively) a permanently reduced sleep duration is associated with hypertension, coronary heart disease, diabetes and obesity [2-4], as well as an increased risk for cognitive impairment [5]. Vice versa, persons with obesity or diabetes show a higher rate of sleep disturbances suggesting a reciprocal interaction between sleep and metabolic state. Although drug treatment represents a quick and easy approach, once discontinued, medication often leads to drug dependence or a return of sleep disturbances [6]. Therefore, a nutritional approach, e.g., changing dietary composition, using certain foods/macronutrients, or changing eating timing offers an interesting way to treat sleep disorders in a natural way.

Accordingly to the two-process model of the sleep regulation, circadian clocks along to the homeostatic regulation contribute to the sleep-wake behavior. Circadian clocks, which synchronize the body’s internal processes with the 24-hour day-night cycle, influencing sleep-wake patterns, hormone release, and overall alertness. In humans, sleep normally occurs during the biological night and is associated with a decrease in body temperature and the synthesis of melatonin [7]. A misalignment of the sleep-wake rhythm from the day-night cycle results in disturbed physiological and hormonal rhythms which are affected at the transcriptional and translational levels [7]. Such separation of behavioral cycle (sleep and/or food intake) from the circadian cycle, which particularly occurs with chronic shift work or acute jet lag, is associated with obesity, type 2 diabetes, and further negative cardiometabolic effects [8, 9]. Moreover, a disturbed sleep-wake rhythm can affect eating behavior and the regulation of appetite [10]. Circadian misalignment of the circadian clock is tightly associated with decreased sleep efficiency and daytime sleepiness [9, 10]. Thus, circadian desynchronization apparently contributes to the bidirectional link between metabolic diseases and poor sleep. In this context, a recent review discusses the role of lifestyle factors such as scheduled eating and sleep as potent tools which can restore disturbed circadian rhythmicity and in this way contribute to the improvement of cardiometabolic outcomes [11].

Time-restricted eating, a dietary strategy characterized by the shortening of daily eating window, was shown to improve both cardiometabolic outcomes and sleep. In published research, the sleep quality was assessed by subjective and subjective approaches showing heterogenous results. For instance, McStay et al. [12] focused their review on the impact of TRE on sleep and refer to a number of TRE studies that assessed sleep quality using the Pittsburgh Sleep Quality Index (PSQI). It has been described that TRE had no effect on sleep quality in either well-sleeping (PSQI <5) or poorly sleeping (PSQI >5) subjects [13-17]. Wilkinson et al. [17] showed a moderate but non-significant decrease in PSQI and a 23% increase in restful sleep compared to baseline using the myCircadianClock (mCC) app in a 12-week, 10-hour TRE intervention. Additionally, trends were observed that led to the hypothesis of longer sleep duration and sleep efficiency due to TRE [17]. However, there was only one TRE study that observed longer sleep duration as a result of the 16-week, 10-h TRE intervention compared with baseline, but no validated study tool was used [18]. Regarding sleep latency and sleep efficiency, Lowe et al. [15] reported a worsening as a result of 8-h TRE in women and men with overweight and obesity. Some TRE trials showed no effect of TRE on sleep latency [13] or sleep efficiency [13, 16].

Notably, sleep restriction and possibly sleep fragmentation are associated with glucose intolerance [19] so that sleep improvement by TRE might reduce the risk of diabetes and cardiovascular diseases [11]. However, due to heterogenous data, further investigations on the effects of TRE on sleep quality and sleep patterns are needed – ideally by objective tools - and general recommendations regarding the optimal eating timing are pending. Therefore, the aim of this study was to compare the effects of early (eTRE) with late (lTRE) time-restricted eating on the sleep quality using subjective and objective methods. This study was conducted in terms of the ChronoFast trial (NCT04351672) [20, 21], where sleep metrics were secondary outcomes.

## 2. Materials and Methods

### 2.1. Study design

This study was conducted within the cohort of the ChronoFast trial, which was carried out from March 2020 to December 2021. ChronoFast was a 10-week randomized, controlled crossover trial which explored the impact of early and late Time-restricted eating on cardiometabolic health outcomes in women with overweight and obesity, without changes in dietary intake and upon nearly unchanged body weight. Participants underwent two dietary interventions: early time-restricted eating (eTRE = eating from 8 am to 4 pm; 14-days) and late time-restricted eating (lTRE = eating from 1 pm to 9 pm; 14-days) under random assignment of the order. Study subjects had a 4-week run in phase and an intermediate 2-week washout phase between dietary interventions. During the TRE-interventions, the study subjects got instructed to eat in the specified eating windows and do not change caloric intake and dietary composition compared to run in phase. In the defined fasting windows, participants were allowed to consume water, tea, black coffee, as well as sugarfree chewing gums or sugarfree softdrinks in small amounts. ChronoFast was approved by Ethical Committee of the University of Potsdam, Germany (EA No. 8/2019, approved on 25.10.2019) and performed in harmony with Helsinki Declaration, 1975. The registration of the ChronoFast trial can be accessed on www.clinicaltrials.gov under the identifier NCT04351672 (registered on April 17, 2020). The study subjects provided their written informed prior to participation in this study. The whole study design and study methods were published previously (21).

### 2.2. Study population

The ChronoFast trial was conducted in a cohort of 31 female study subjects aged 18 to 70 years, a BMI between 25 and 35 kg/m^2^, and a Pittsburgh Sleep Quality Index (PSQI) <10, as a guideline for a good sleep quality prior study start. Participants were from Germany, recruited in the area of Berlin-Brandenburg. Recruitment was performed by personal interview, posters, flyers, newspaper advertisements and selected ads websites. The study was conducted at the German Institute of Human Nutrition.

Prior to participation, all voluntary enrolled subjects gave their written consent after being informed. The trial was approved by Ethical Committee of the University of Potsdam, Germany (EA No. 8/2019) and conducted in line with the Helsinki Declaration, 1975. Exclusion criteria were conditions interfering with variables of interest, e.g. shift work, traveling over different time zones and recent weight changes (>5 %). Also, several diseases, a.o., diabetes type 1 or 2, severe kidney, liver, psychiatric diseases, as well as various medication led to exclusion. A detailed listing of exclusion criteria can be viewed in the previously published study protocol (21).

### 1.1. Data collection

#### 1.1.1. Dietary intake and composition

Dietary intake, e.g., energy intake and amounts of macronutrients, was accessed during run in phase (14-days), TRE-interventions (each 14-days) and in the intermediate washout phase (3-days; 2 weekdays + 1 weekend day). Participants were enabled to record food intake, eating times and weight via paper-based documentation or with the smartphone FDDB Extender app (FDDB Internetportale GmbH, Bremen, Germany), which we already highlighted as a valid and practical research tool for capturing energy and macronutrient intake (22). Previously handwritten logs were transferred into a new generated FDDB participant-account by study staff and therefore got analysed based on FDDB database (Fddb Internetportale GmbH, https://fddb.info/).

#### 1.1.2. Pittsburgh sleep quality index and self-evaluation of sleep quality

The PSQI was recorded during pre-screening (inclusion criteria: PSQI <10) and at every visit pre and post TRE-intervention phases to assess changes in subjective sleep quality. The PSQI questionnaire contained 19 items on sleep duration, sleep latency, sleep efficiency, subjective sleep quality and sleep disturbances, as well as the use of sleep medications, based on which a global evaluation of sleep quality was made (23). Another subjective tool for further insights in perceived sleep quality was a self-evaluation. Thereby, participants used a school grade system (1 = “very good” to 5 = “very bad”) to evaluate the past night in the next morning.

#### 1.1.3. Actigraphy and sleep logs

Objective Sleep quality was assessed with an accelerometer (ActiGraph wGT3X-BT, ActiGraph) in the Run-in phase (14-days) and both TRE-interventions (each 14-days), which participants wore around their non-dominant wrist. The device was worn constantly. Removal of the device was only required if the study participants were swimming or taking a sauna. In the case of removal, the time and duration were determined paper-based by the study participant. Moreover, the timing of sleep, e.g., bed time and wake-up time, was logged daily by study participants simultaneously to the wearing of ActiGraph device. In addition, the activity records and documented sleep times were used to calculate total sleep duration, time to fall asleep in minutes, sleep efficiency in percent, as well as the Sleep Fragmentation Index (SFI) as an indicator of restlessness during the sleep period (<u>24</u>). The calculation performed with the related ActiLife software version 6.13.4 (ActiGraph, Pensacola, FL, USA) using the Sadeh algorithm.

Logged sleeping times for calculation of mentioned parameters were transferred manually by study staff into the ActiLife software. Days with a wear time of less than 50% and days on which the Actigraph was handed out to the study subjects or returned were not included into analysis.

#### 1.1.4. Hunger and satiety scores

The assessment of the states of hunger and satiety was performed with a questionnaire based on four questions concerning 1. the desire to eat something now, 2. the current feeling of hunger, 3. the current feeling of satiety, and 4. the amount of food that can currently be consumed. Each question was answered on the last day of the TRE intervention, both in the morning at 8 am. and in the evening at 8 pm, by drawing a line on a visual analogue scale (VAS) of ten centimetres (0 = “not at applicable at all” to 10 = “fully applicable”).

### 1.2. Statistical analysis

For Power calculation we used the software G-Power v.3.1 (25), whereas the for the primary end point was defined as change in insulin sensitivity and was based on our previous study where the difference of insulin sensitivity in meal tolerance test (morning vs. evening) was examined in pre-diabetic subjects (26). The present study was planned for a sample size of 30 and therefore powered for detection of an 0.53 effect size, a significance level 0.05 and 80% statistical power. Due to minimization method, using MinimPy software (27), an allocation of study participants to study arms based on their BMI and age was performed.

Statistical analysis was performed using SPSS 25.0 software (IBM, Chicago, IL, United States). Mean values of the collected data and the standard errors of the mean (SEM) are shown. The distribution of the data was determined using the Shapiro-Wilk test. A parametric Student’s t-test (paired) was performed for normally distributed data. Non-normally distributed data was log-transformed and re-assessed for normality. In case the assumption of a normally distributed sample for the dependent samples t-test were not fulfilled even after a log-transformation, a non-parametric Wilcoxon sign rank test was chosen for analysis. To compare the effects of the TRE interventions on PSQI, which was determined before and after each intervention, with each other, an individual delta percentage value (Δ%) was calculated for each TRE intervention and each participant as follows: (post-intervention ± pre-intervention)/pre-intervention*100. With determined Δ%-values the statistical analysis was then carried out as described above. For the definition of statistical significance, a p value <0.05 was defined. The correlation was tested using a Pearson test. All graphs for data were visualized with GraphPad Prism software version 5.0 (GraphPad Prism Inc, La Jolla, CA, USA).

## 2. Results

A total of 90 subjects were pre-screened by telephone through study nurses and with questionnaires, whereas 36 of pre-screened subjects met eligibility criteria and underwent a personal screening in study centre. 31 of them enrolled run-in phase and got randomized. Thereby, 15 participants were allocated to group A (1. eTRE – 2. lTRE) and 16 participants to group B (1. lTRE – 2. eTRE). In totally all 31 subjects completed the study and the data of them was analyzed.

**Figure 1.**
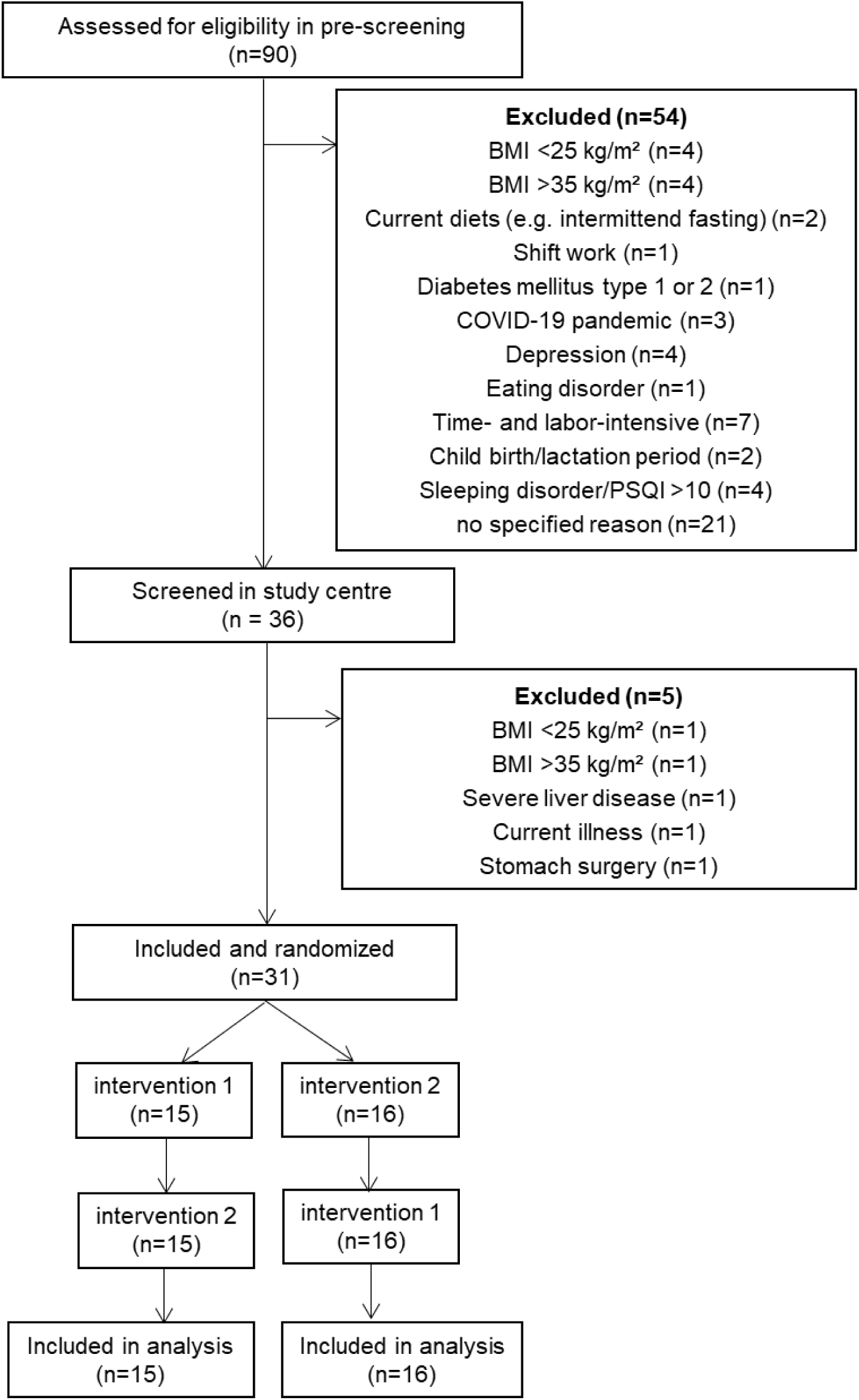
Flowchart of participant eligibility.

### 2.1. Clinical characteristics of participants

The baseline characteristics of study population at study entry are shown in **Table 1**. A total of 31 participants completed the ChronoFast study. Subjects were 58.39 ± 1.53 years old, and moderately overweight with a BMI of 30.47 ± 0.52 kg/m^2^. 18 participants showed a normal glucose tolerance, whereas 13 participants were pre-diabetic with an impaired fasting glucose and/or an impaired glucose tolerance. An average HbA1c of 5.48 ± 0.06 % was determined. The adherence to timed eTRE and lTRE eating windows was very high with over 96 % in both interventions and therefore eating time was reduced from 12:06 ± 0:17 h in Run-in to 7:09 ± 0:05 h in eTRE-intervention and 6:57 ± 0:09 h in lTRE-intervention. A spontaneous weight loss, but under the border of feared influence on metabolism (< 5 %), occurred in both TRE-interventions (eTRE: -1.08 kg, p = 9.12×10^-8^; lTRE: - 0.44 kg, p = 0.007) and therefore related changes of BMI were determined (eTRE: -0.45 kg/m^2^, p = 5.36×10^-9^; lTRE: -0.12 kg/m^2^, p = 0.014). Body composition changed in eTRE with decreases of total body fat (-0.75 kg, p = 0.001), whereas lean mass remained unchanged within both TRE interventions. While energy intake was reduced within eTRE-intervention (eTRE: -167.24 kcal, p =2.38×10^-4^), macronutrient composition (EN%, Energy percentage of each macronutrient in relation to total calorie intake) remained unchanged in both TRE-interventions compared to run-in phase, whereby two participants was excluded in calorie and macronutrient analysis due to observed underreporting. Physical activity in TRE intervention periods stayed stable as before shown in run-in period. Participants did not change their habitual physical activity, assessed by the parameter metabolic equivalent of task (MET) and percentages of light, moderate and sedentary activity.

**Table 1.** Baseline characteristics of study population.

|  |  |
| --- | --- |
| Females [n] | 31 |
| Age | 58.39 ± 1.53 |
| Randomization |  |
| A. eTRE – ITRE | 15 |
| B. ITRE – eTRE | 16 |
| Glycemic state [n] |  |
| NGT | 18 |
| IFG/IGT | 13 |
| Chronotype [n] |  |
| early | 18 |
| normal | 11 |
| late | 2 |
| PSQI | 5.68 ± 0.37 |
| Weight [kg] | 82.53 ± 1.51 |
| BMI [kg/m<sup>2</sup>] | 30.47 ± 0.52 |
| Hip circumference [cm] | 111.74 ± 1.08 |
| Waist circumference [cm] | 99.22 ± 1.60 |
| Waist-to-hip-ratio | 0.89 ± 0.01 |
| Fat mass [kg] | 34.76 ± 1.11 |
| Fat-free mass [kg] | 47.48 ± 0.84 |
| Systolic blood pressure [mmHg] | 121.39 ± 2.62 |
| Diastolic blood pressure [mmHg] | 73.16 ± 1.16 |

All data are shown as mean ± SEM (n = 31). Abbreviations: eTRE, early Time-restricted eating; ITRE, late Time-restricted eating; NGT, normal glucose tolerance; IFT, impaired fasting glucose; IGT, impaired glucose tolerance; PSQI, Pittsburgh Sleep Quality Index; BMI, Body-Mass-Index.

### 2.2. Subjective Sleep quality

To investigate effects of eTRE and lTRE on subjective sleep quality we used PSQI tool. In pre-post-comparison, no significant changes in subjective sleep quality were found during eTRE and lTRE **(Figure 1A)**. Also, no differences between TRE interventions, calculated as Δ%, were observed. Moreover, study subjects self-reported their sleep quality to be the same as before the TRE interventions **(Figure 1B).**

**Figure 1.**
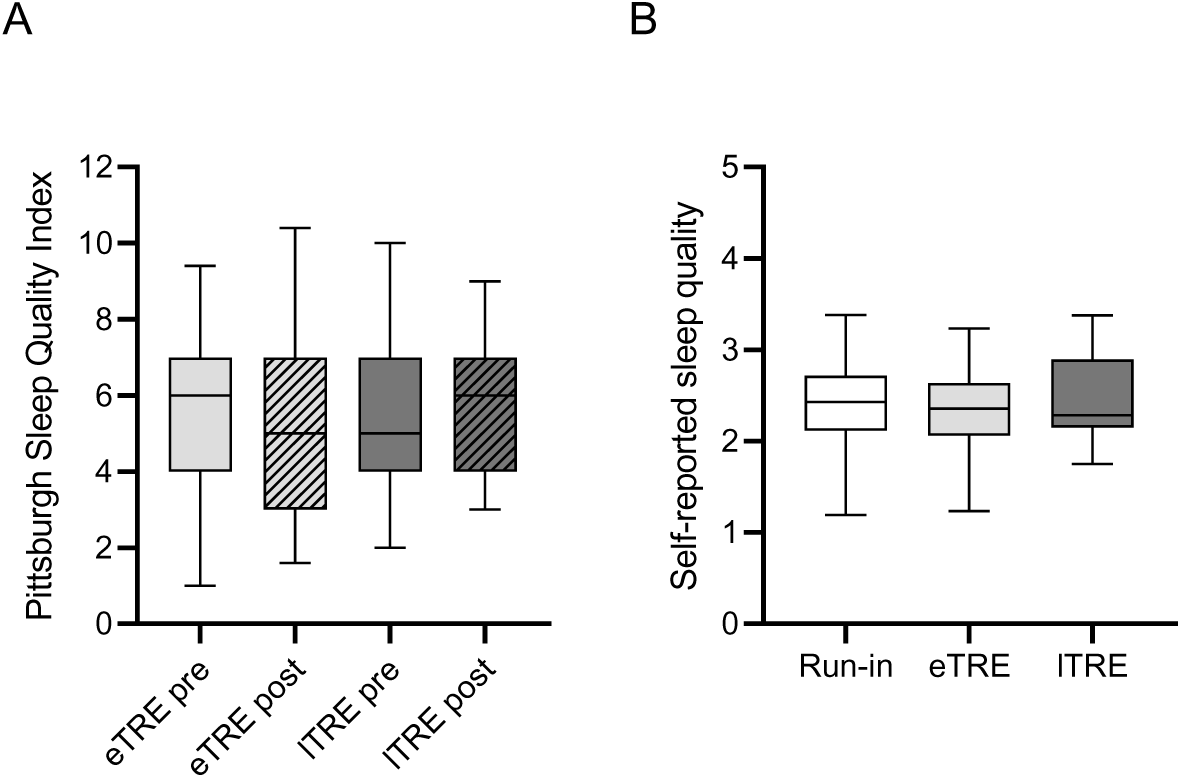
Effects of eTRE and lTRE on subjective sleep quality. The Pittsburgh Sleep Quality Index (A) pre and post eTRE and lTRE, as well as the self-reported sleep quality during the run-in and TRE-intervention phases (B) is presented. PSQI-data is shown as Boxplot with whiskers displaying 5% – 95% confidence interval (n = 31). Self-reported sleep quality-data is presented as Boxplot with whiskers displaying 5% – 95% confidence interval (Run-in: n = 30; eTRE: n =30; lTRE: n = 29). *p < 0.05. Missing data of self-reported sleep quality is based on an incorrectly/not completed sleep protocol.

### 2.3. Objective Sleep Quality

Objective Sleep Quality and several sleep parameters were assessed by ActiGraph wGT3X-BT. Therefore, sleep latency remained unchanged within both TRE-interventions **(Figure 2A**). Significant differences between phases were not observed. Total sleep time, showed to be extended from 394.12 ± 7.87 min in the run-in phase to 406.10 ± 7.88 min in the lTRE intervention, but differences were not significant in statistical analysis (p = 0.101). A total sleep time about 398.17 ± 7.77 min in eTRE was almost similar to run-in phase (**Figure 2C**). Sleep efficiency, calculated as percentage of time sleeping of total spend time in bed, was increased significantly in eTRE intervention compared to run-in phase (p = 0.047) (**Figure 2B**). Meanwhile Sleep Fragmentation Index (SFI), a parameter for restless sleep, was decreased in eTRE intervention compared to run-in phase and led to hypothesis of calmer sleep through eTRE (p = 0.029) **(Figure 2D**).

**Figure 2.**
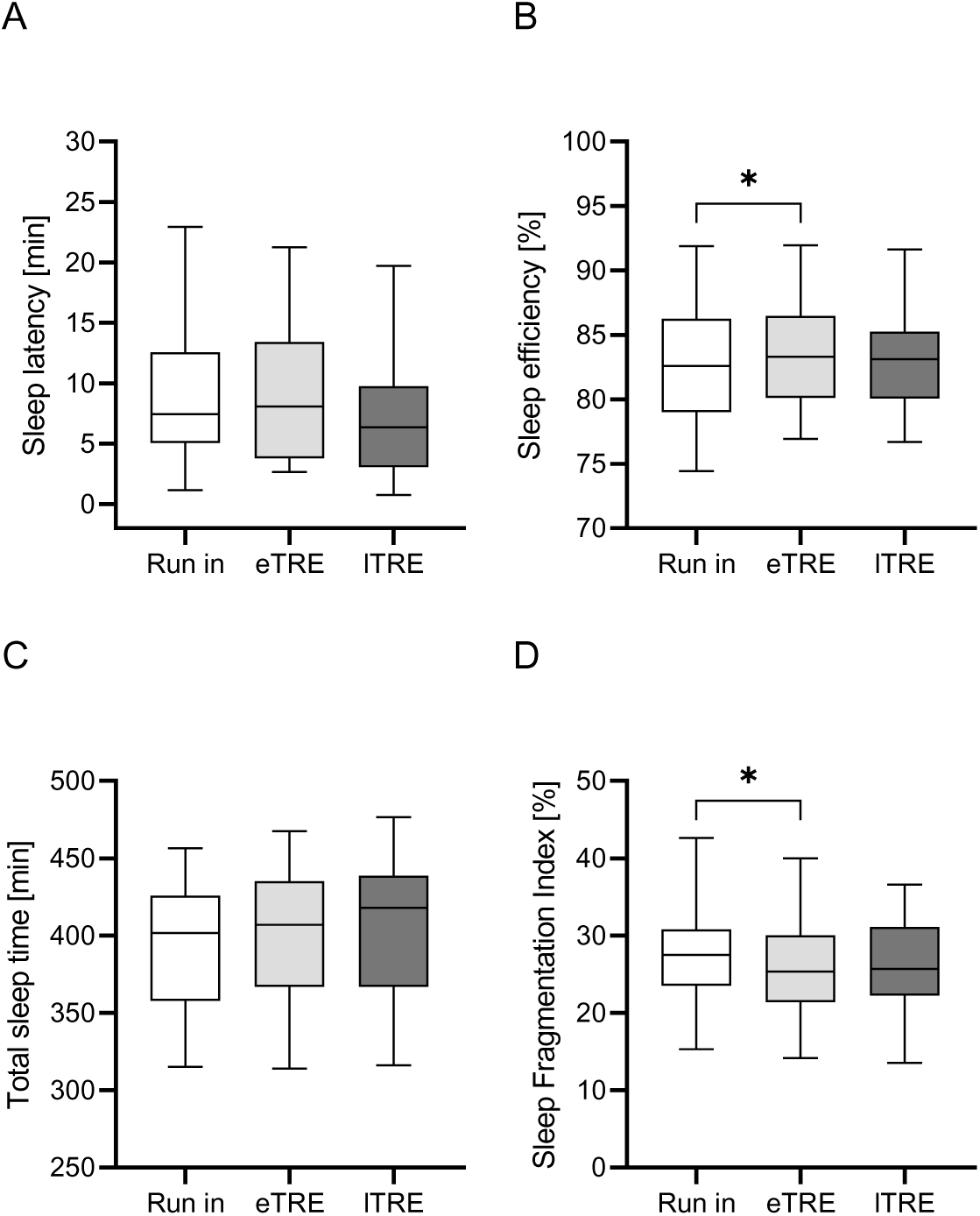
Effects of eTRE and lTRE on objective sleep metrics. Shown are the parameters sleep latency (A), sleep efficiency (B), total sleep time (C) and Sleep Fragmentation Index (D) in the run-in phase, eTRE- and lTRE-intervention, assessed via actigraphy. Data for Sleep-Latency is shown as Boxplot with whiskers displaying 5% – 95% confidence interval (Run-in: n = 27; eTRE: n = 29, lTRE: n = 30). Data for sleep-efficiency, total sleep time and Sleep Fragmentation Index is shown as Boxplot with whiskers displaying 5% – 95% confidence interval (Run-in: n = 28, eTRE and lTRE: n = 31). Measurement errors for three participants in run-in occurred due to defect accelerometers. Further for Sleep Latency one more value in Run-in, two values in eTRE and one value in lTRE were determined as outliner and therefore were excluded from analysis. *p < 0.05.

### 2.4. Hunger and Satiety

Due to the restriction of food, we assumed a correlation of hunger and satiety with sleep and therefore analysed the parameters of Hunger and Satiety with a VAS. VAS data showed no differences of hunger and satiety parameters in the evening of the last intervention day.

**Figure 3.**
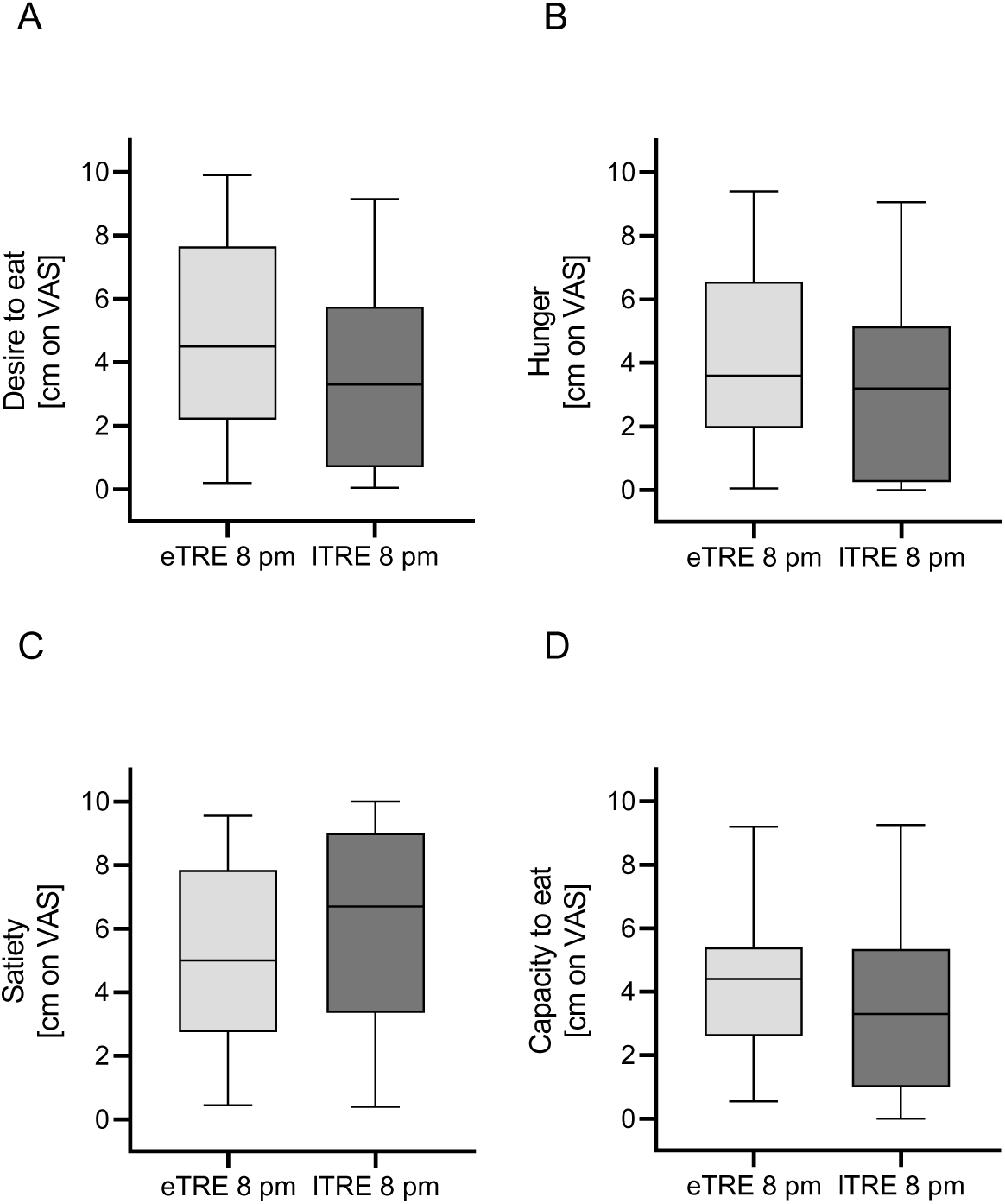
Effects of eTRE and lTRE on hunger and satiety scores. The Desire to eat (A), the feeling of hunger (B) and satiety (C), as well as the capacity to eat (D) at the end of the eTRE- and lTRE-intervention (last day 8 pm), assessed on a visual analog scale (VAS) is shown. Data for desire to eat, satiety and capacity to eat is presented as Boxplot with whiskers displaying 5% – 95% confidence interval (eTRE n = 29; lTRE n = 29). Data for hunger is shown as Boxplot with whiskers displaying 5% – 95% confidence interval (eTRE n = 29; lTRE n = 28). Data sets of two participants at eTRE and two participants at lTRE were excluded in the analysis due to misunderstandings in completing the VAS. Further one participant forgot to fill out a hunger-value in lTRE. *p < 0.05.

To examine the relationship between the parameters of hunger and satiety with sleep quality, a Pearson correlation test was performed for both intervention phases. Sleep efficiency and SFI were used as parameters of sleep quality. For the parameter of the feeling of hunger, there was only a very small, but non-significant, negative correlation during the lTRE intervention phase in connection with the SFI (r = -0.060; p = 0.772). All other tests also showed no significant results.

## 3. Discussion

### 3.1. Early Time-restricted eating leads to more effective and restful sleep

A PSQI under 5 is associated with a good sleep quality (13). In ChronoFast study the PSQI at the baseline was 5.68 ± 0.37. Thus, previous reference values lead to the classification of a Quelle einfügen moderate sleep quality prior study start. With regard to the examination of PSQI, we neither observed positive nor negative effects on objective sleep quality due to eTRE or lTRE. This finding is consistent with those of other studies (14-18).

In the review about Sleep and TRE by McStay et al. (13) minimal losses of body weight under 2 to 3 % were discussed as a reason for absence of positive effects on sleep quality. This assumption could be also a reason for absence of effects in ChronoFast study. Although spontaneous weight loss occurred because of restriction of eating window, weight loss was under 5 % which we expected to have a metabolic effect. In this context McStay et al. (13) refer to an effect starting from 5 % weight loss. Moreover, Wilkinson et al. (18) did not observe significant effects in a 12-week 10h-TRE study with PSQI-Tool, but with an app-based questionnaire assessment method on daily sleep quality. Also, Kesztyüs et al. (28) observed an improvement of sleep quality using a VAS-Tool, but not with PSQI-Tool. McStay et al. (13) highlighted memory bias and low measurement recoveries to be lower in VAS- or myCircadianClock-app-based assessment. Due to this, we additionally measured objective sleep quality by self-evaluation the ChronoFast study, but again no effects of eTRE and lTRE on self-evaluated sleep quality occurred.

Analysis of objective sleep quality was performed with accelerometer as described above. Literature refers to an optimal sleep duration between 7 to 8 h (11). In the ChronoFast study, no change in sleep duration was observed within both TRE-interventions. This result is only not supported by the study of Gill and Panda (19), although McStay et al. (13) caution interpretation of the results referring to an unvalidated study tool in their review on the effects of TRE on sleep.

Regarding sleep latency we neither found changes in comparison to run-in phase, nor between eTRE- and lTRE-interventions. However, we found a positive effect of eTRE on sleep efficiency. In contrast, another study of Lowe et al. (16) reported a worsening of sleep efficiency in TRE with a window of caloric intake between 12 am and 8 pm which is very similar to the lTRE of the ChronoFast study. Besides sleep efficiency, the eTRE intervention also led to a significant improvement of SFI. Chung et al. (29) examined how food intake 3 h before bedtime affected sleep quality. The results showed an association with nocturnal awakening, but no increase in time to fall asleep or shortened sleep duration. This suggests that lTRE has a less positive effect on sleep than eTRE. Therefore, we assume that eTRE led to calmer sleep with less interruptions.

### 3.2. Improved sleep metrics are independently from hunger- and satiety-feelings

A recently emerged hypothesis, discusses insufficient sleep being a promotor for energy intake based on the assumption of eating in absence of hunger (30). Therefore, it has been discussed that more wake-time opens more possibilities for food intake (30). Based on this and current research on interactions between dietary modifications and sleep and alterations in appetite due to restricted sleep (31), we investigated on accompanying changes of hunger/satiety due to TRE in ChronoFast study.

Thus, a recently published crossover study investigated on the effects of lTRE as a promotor for a positive energy balance and showed increased self-reported hunger, as well as changed hormone levels. The authors reported decreased waketime 24-h-leptin levels and a higher (acetylated) ghrelin:leptin ratio, which is associated with hunger, after lTRE-diet. However, the hypothesis of an impact of appetite on the sleep parameter, a.o., total sleep time, sleep efficiency, wake-time, was not confirmed (31). Lower hunger in participants who loaded main calories in the morning compared to participants who consumed most of their calories in the evening was also shown in another crossover study and led to hypothesis that early eating could improve compliance to a weight loss diet (32). Our present study agrees with this, because isocaloric early eating showed to be more difficult to perform without spontaneous weight loss. Thus, participants in ChronoFast study lost 0.64 kg more weight in eTRE than in lTRE intervention. Therefore, we hypothetized nutrient intake in the morning containing higher amounts of fiber and therefore making participants more saturated. In addition, we assumed snacking, which is associated with higher amounts of short-chain carbohydrates, being more present in the evening and could lead to satisfaction of appetite, but not nutrient-hunger. However, no changes in macronutrient intake due to TRE were not found in both interventions and the stated hypothesis had to be rejected for now. Further investigation on this has to be done through assessment of nutrient pattern and food cravings in the morning and evening present an interesting research gap. Thus, higher satiety and less food cravings could lead to a higher success of TRE and a higher success, when weight loss is targeted. In ChronoFast study, participants were requested not to change dietary composition. If participants would have been allowed to eat as they wished, outcomes for the feeling of hunger and satiety may be different.

All in all, we assume that changes of hunger and satiety, as well as changes of sleep are not directly associated to each other, but both influenced independently through TRE. However, we have shown that eTRE led to a higher sleep efficiency and less restless sleep.

## Limitations

For the evaluation of the results it is important to mention that the measurements of the actigraph were analyzed with the Sadeh algorithm. This algorithm had been developed using data from young adults aged 20-25 years and children (33). Since the age of the subjects in this study ranged from 18 to 70 years, it is necessary to consider whether the use of the Cole Kripke algorithm, which is based on data from adults of different ages (34), would produce different results. Furthermore, it has to be considered that due to the manual logging a recall bias cannot be excluded if the sleep logs were not filled in exactly at the time of going to bed. Moreover, a potential influence of daytime sleep on nighttime sleep was not included in this analysis and might have had an impact as well.

## Conclusions

Due to our research we have shown eTRE to have positive effects on objective metrics of sleep. In ChronoFast study, a bad sleep quality and sleep disorders prior to study start, were defined as exclusion criteria. However, we assume that TRE could be more beneficial for patients with a bad sleep quality in the baseline, although this effect was not confirmed in a previous study in study subjects with sleep disorders (14). Further investigations have to focus on the effect of hunger on the compliance of TRE which is a main indicator for the success of a diet.

## Data Availability

All data produced in the present study are available upon reasonable request to the authors

## Author Contribution

Beeke Peters: investigation, data collection, formal analysis, writing original draft, review and editing. Jennifer Jokisch: data collection, formal analysis. Julia Schwarz, Bettina Schuppelius: investigation, data collection, formal analysis. Nico Steckhan: formal analysis, writing-review and editing. Andreas Michalsen, Achim Kramer: conceptualization, writing-review and editing. Olga Pivovarova-Ramich: conceptualization, writing original draft, review and editing.

## Acknowledgements

We acknowledge all study participants for their cooperation. We gratefully thank Manuela Bergmann for the support in the clinical trial conduction, Juliane Roeder, Melanie Hannemann, and Lothar Napieralski for their work with study subjects, as well as Katja Treu and Christiana Gerbracht for the help in the preparation of nutritional counselling and dietary record analysis.

## Supplementary Material

### Supplement 1. Sleep metrics

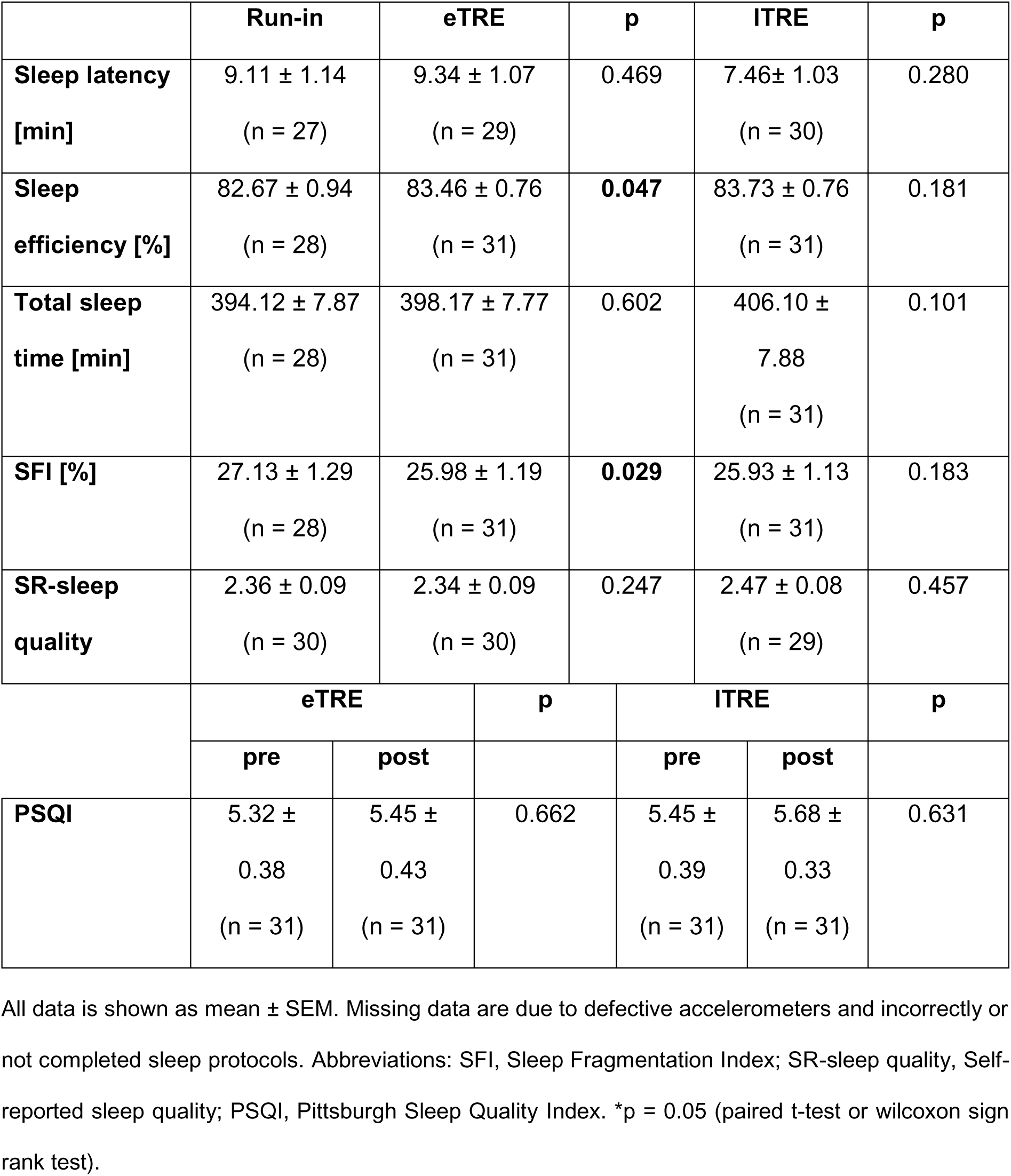

### Supplement 2. Hunger and satiety scores

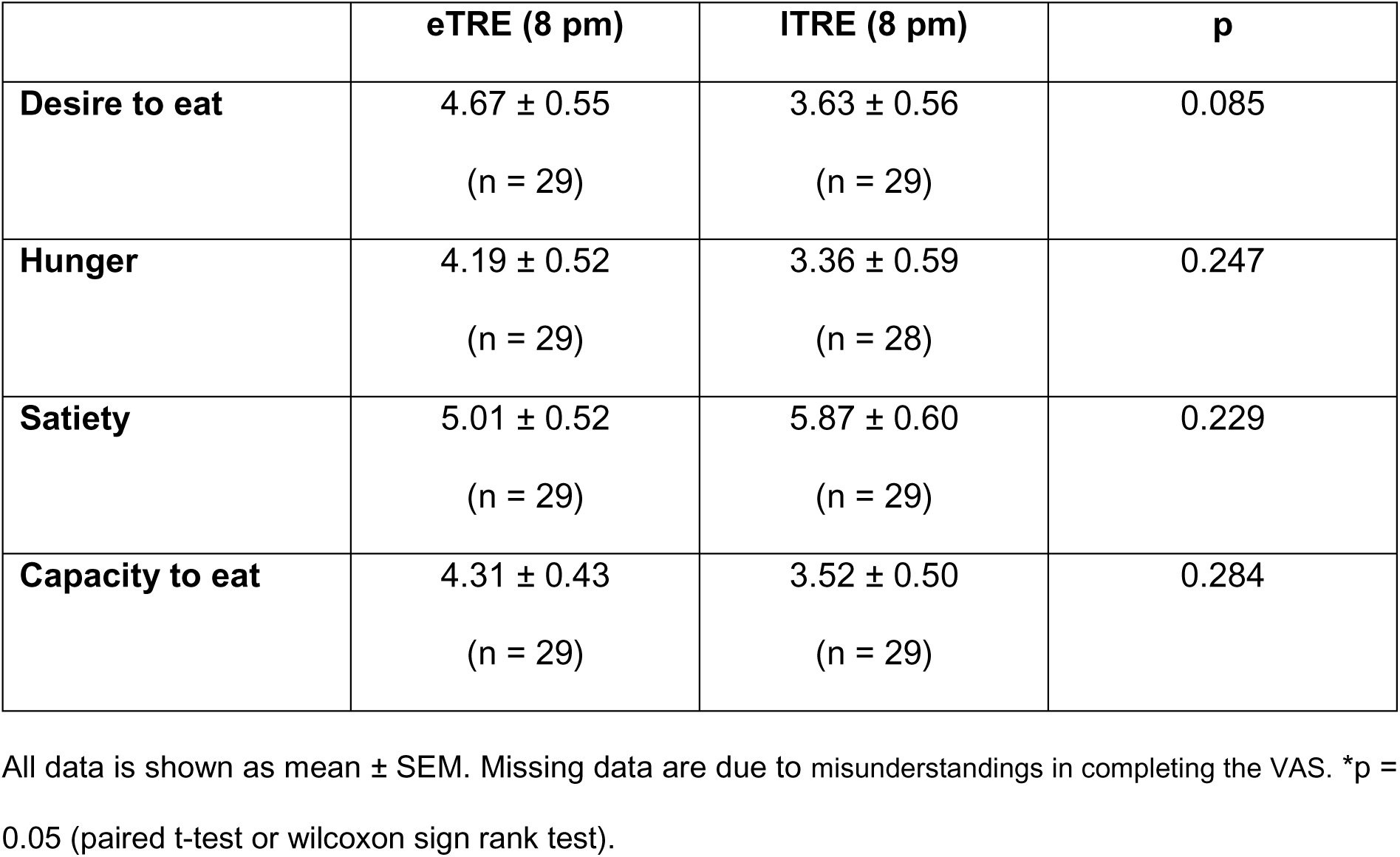

**Supplement 2. Hunger and satiety scores.**
|  | **eTRE (8 pm)** | **ITRE (8 pm)** | **p** |
| --- | --- | --- | --- |
| **Desire to eat** | 4.67 ± 0.55<br>(n = 29) | 3.63 ± 0.56<br>(n = 29) | 0.085 |
| **Hunger** | 4.19 ± 0.52<br>(n = 29) | 3.36 ± 0.59<br>(n = 28) | 0.247 |
| **Satiety** | 5.01 ± 0.52<br>(n = 29) | 5.87 ± 0.60<br>(n = 29) | 0.229 |
| **Capacity to eat** | 4.31 ± 0.43<br>(n = 29) | 3.52 ± 0.50<br>(n = 29) | 0.284 |

All data is shown as mean ± SEM. Missing data are due to misunderstandings in completing the VAS. \*p = 0.05 (paired t-test or wilcoxon sign rank test).

### Supplement 3: Correlations of hunger and satiety parameters

|  |  |  | eTRE |  | ITRE |  |
| --- | --- | --- | --- | --- | --- | --- |
|  |  |  | Hunger | Satiety | Hunger | Satiety |
| eTRE | SE | r | 0.065 | 0.195 |  |  |
| | | $\sigma$ | 0.748 | 0.329 | | |
|  | SFI | r | 0.071 | -0.064 |  |  |
| | | $\sigma$ | 0.726 | 0.749 | | |
| ITRE | SE | r |  |  | 0.131 | -0.082 |
| | | $\sigma$ | | | 0.525 | 0.685 |
|  | SFI | r |  |  | -0.060 | 0.005 |
| | | $\sigma$ | | | 0.772 | 0.979 |

Abbreviations: r, Pearson correlation; SE, Sleep efficiency; SFI, Sleep Fragmentation Index.

### Supplement 4. Correlations of sleep parameters

**Supplement 4. Correlations of sleep parameters.**
|  |  |  | eTRE | ITRE |
| --- | --- | --- | --- | --- |
|  |  |  | PSQI |  |
| eTRE | SE | r | -0.188 |  |
|  |  | σ | 0.312 |  |
|  | SFI | r | 0.224 |  |
|  |  | σ | 0.226 |  |
| ITRE | SE | r |  | -0.001 |
|  |  | σ |  | 0.997 |
|  | SFI | r |  | 0.150 |
|  |  | σ |  | 0.421 |

Abbreviations: r, Pearson correlation; SE, Sleep efficiency; SFI, Sleep Fragmentation Index; PSQI, Pittsburgh Sleep
Quality Index.

## Literature

1. Chattu VK, Manzar MD, Kumary S, Burman D, Spence DW, Pandi-Perumal SR. The Global Problem of Insufficient Sleep and Its Serious Public Health Implications. Healthcare (Basel). 2018;7(1).

2. Cappuccio FP, Stranges S, Kandala NB, Miller MA, Taggart FM, Kumari M, et al. Gender-specific associations of short sleep duration with prevalent and incident hypertension: the Whitehall II Study. Hypertension. 2007;50(4):693–700.

3. Hasler G, Buysse DJ, Klaghofer R, Gamma A, Ajdacic V, Eich D, et al. The association between short sleep duration and obesity in young adults: a 13-year prospective study. Sleep. 2004;27(4):661–6.

4. Svensson AK, Svensson T, Kitlinski M, Almgren P, Engström G, Nilsson PM, et al. Incident diabetes mellitus may explain the association between sleep duration and incident coronary heart disease. Diabetologia. 2018;61(2):331–41.

5. Fernandez-Mendoza J, He F, Puzino K, Amatrudo G, Calhoun S, Liao D, et al. Insomnia with objective short sleep duration is associated with cognitive impairment: a first look at cardiometabolic contributors to brain health. Sleep. 2021;44(1).

6. Victorri-Vigneau C, Dailly E, Veyrac G, Jolliet P. Evidence of zolpidem abuse and dependence: results of the French Centre for Evaluation and Information on Pharmacodependence (CEIP) network survey. Br J Clin Pharmacol. 2007;64(2):198–209.

7. Archer SN, Laing EE, Moller-Levet CS, van der Veen DR, Bucca G, Lazar AS, et al. Mistimed sleep disrupts circadian regulation of the human transcriptome. Proc Natl Acad Sci U S A. 2014;111(6):E682–91.

8. Antunes LC, Levandovski R, Dantas G, Caumo W, Hidalgo MP. Obesity and shift work: chronobiological aspects. Nutr Res Rev. 2010;23(1):155–68.

9. Scheer FA, Hilton MF, Mantzoros CS, Shea SA. Adverse metabolic and cardiovascular consequences of circadian misalignment. Proc Natl Acad Sci U S A. 2009;106(11):4453–8.

10. Baron KG, Reid KJ. Circadian misalignment and health. Int Rev Psychiatry. 2014;26(2):139–54.

11. Sheikh-Ali M, Maharaj J. Circadian clock desynchronisation and metabolic syndrome. Postgrad Med J. 2014;90(1066):461-6.

12. Gupta CC, Vincent GE, Coates AM, Khalesi S, Irwin C, Dorrian J, et al. A Time to Rest, a Time to Dine: Sleep, Time-Restricted Eating, and Cardiometabolic Health. Nutrients. 2022;14(3).

13. McStay M, Gabel K, Cienfuegos S, Ezpeleta M, Lin S, Varady KA. Intermittent Fasting and Sleep: A Review of Human Trials. Nutrients. 2021;13(10).

14. Cienfuegos S, Gabel K, Kalam F, Ezpeleta M, Pavlou V, Lin S, et al. The effect of 4-h versus 6-h time restricted feeding on sleep quality, duration, insomnia severity and obstructive sleep apnea in adults with obesity. Nutr Health. 2022;28(1):5–11.

15. Gabel K, Hoddy KK, Burgess HJ, Varady KA. Effect of 8-h time-restricted feeding on sleep quality and duration in adults with obesity. Appl Physiol Nutr Metab. 2019;44(8):903–6.

16. Lowe DA, Wu N, Rohdin-Bibby L, Moore AH, Kelly N, Liu YE, et al. Effects of Time-Restricted Eating on Weight Loss and Other Metabolic Parameters in Women and Men With Overweight and Obesity: The TREAT Randomized Clinical Trial. JAMA Intern Med. 2020;180(11):1491–9.

17. Parr EB, Devlin BL, Lim KHC, Moresi LNZ, Geils C, Brennan L, et al. Time-Restricted Eating as a Nutrition Strategy for Individuals with Type 2 Diabetes: A Feasibility Study. Nutrients. 2020;12(11).

18. Wilkinson MJ, Manoogian ENC, Zadourian A, Lo H, Fakhouri S, Shoghi A, et al. Ten-Hour Time-Restricted Eating Reduces Weight, Blood Pressure, and Atherogenic Lipids in Patients with Metabolic Syndrome. Cell Metab. 2020;31(1):92–104 e5.

19. Gill S, Panda S. A Smartphone App Reveals Erratic Diurnal Eating Patterns in Humans that Can Be Modulated for Health Benefits. Cell Metab. 2015;22(5):789–98.

20. Reutrakul S, Van Cauter E. Sleep influences on obesity, insulin resistance, and risk of type 2 diabetes. Metabolism. 2018;84:56–66.

21. Peters B, Koppold-Liebscher DA, Schuppelius B, Steckhan N, Pfeiffer AFH, Kramer A, et al. Effects of Early vs. Late Time-Restricted Eating on Cardiometabolic Health, Inflammation, and Sleep in Overweight and Obese Women: A Study Protocol for the ChronoFast Trial. Front Nutr. 2021;8:765543.

22. Baum Martinez I, Peters B, Schwarz J, Schuppelius B, Steckhan N, Koppold-Liebscher DA, et al. Validation of a Smartphone Application for the Assessment of Dietary Compliance in an Intermittent Fasting Trial. Nutrients. 2022;14(18).

23. Buysse DJ, Reynolds CF, 3rd, Monk TH, Berman SR, Kupfer DJ. The Pittsburgh Sleep Quality Index: a new instrument for psychiatric practice and research. Psychiatry Res. 1989;28(2):193–213.

24. Knutson KL, Van Cauter E, Zee P, Liu K, Lauderdale DS. Cross-sectional associations between measures of sleep and markers of glucose metabolism among subjects with and without diabetes: the Coronary Artery Risk Development in Young Adults (CARDIA) Sleep Study. Diabetes Care. 2011;34(5):1171–6.

25. Faul F, Erdfelder E, Lang AG, Buchner A. G*Power 3: a flexible statistical power analysis program for the social, behavioral, and biomedical sciences. Behav Res Methods. 2007;39(2):175–91.

26. Kessler K, Hornemann S, Petzke KJ, Kemper M, Kramer A, Pfeiffer AF, et al. The effect of diurnal distribution of carbohydrates and fat on glycaemic control in humans: a randomized controlled trial. Sci Rep. 2017;7:44170.

27. Saghaei M. An overview of randomization and minimization programs for randomized clinical trials. J Med Signals Sens. 2011;1(1):55–61.

28. Kesztyus D, Fuchs M, Cermak P, Kesztyus T. Associations of time-restricted eating with health-related quality of life and sleep in adults: a secondary analysis of two pre-post pilot studies. BMC Nutr. 2020;6(1):76.

29. Chung N, Bin YS, Cistulli PA, Chow CM. Does the Proximity of Meals to Bedtime Influence the Sleep of Young Adults? A Cross-Sectional Survey of University Students. Int J Environ Res Public Health. 2020;17(8).

30. Chaput J-P. Does sleep restriction increase eating in the absence of hunger? Maybe! The American Journal of Clinical Nutrition. 2021;114(4):1270–1.

31. Vujovic N, Piron MJ, Qian J, Chellappa SL, Nedeltcheva A, Barr D, et al. Late isocaloric eating increases hunger, decreases energy expenditure, and modifies metabolic pathways in adults with overweight and obesity. Cell Metab. 2022;34(10):1486–98 e7.

32. Ruddick-Collins LC, Morgan PJ, Fyfe CL, Filipe JAN, Horgan GW, Westerterp KR, et al. Timing of daily calorie loading affects appetite and hunger responses without changes in energy metabolism in healthy subjects with obesity. Cell Metab. 2022;34(10):1472–85.e6.

33. Sadeh A, Sharkey KM, Carskadon MA. Activity-based sleep-wake identification: an empirical test of methodological issues. Sleep. 1994;17(3):201–7.

34. Cole RJ, Kripke DF, Gruen W, Mullaney DJ, Gillin JC. Automatic sleep/wake identification from wrist activity. Sleep. 1992;15(5):461–9.

